# Identification and Analysis of Novel Synovial Tissue-based Biomarkers and Interacting Pathways for Rheumatoid Arthritis

**DOI:** 10.1101/2021.03.19.21253995

**Authors:** Paridhi Latawa

**Affiliations:** Liberal Arts and Science Academy (LASA), Austin, Texas, USA

## Abstract

Rheumatoid arthritis (RA) is an inflammatory disorder with autoimmune pathogenesis characterized by the immune system attacking the synovium. It is a clinically heterogeneous disease that affects approximately 1.2 million Americans and 20 million people worldwide. It is advantageous to diagnose RA before extensive erosion as treatments are more effective at early stages. RA treatments have made notable progress, yet a significant number of patients still fail to respond to current medication and most of these come with harmful side effects. While the mechanistic reason for such failure rates remains unknown, the cellular and molecular signatures in the synovial tissues of patients with RA are likely to play a role in the variable treatment response and heterogeneous clinical evolution. While blood-based criteria are currently employed for diagnostics and treatments. such serologic parameters do not necessarily reflect biological actions in the target tissue of the patient and are relatively nonspecific to RA. Synovial tissue-based biomarkers are especially attractive as they can provide a confirmed diagnosis for RA. The shortage of accurate synovial tissue-based identifiers for RA diagnosis encouraged this research. This study analyzed data from several gene expression studies for differentially expressed genes in donor synovial tissue. Bioinformatics tools were used to construct and analyse protein interaction networks. Analysis deduced that regulating hematopoietic stem cell migration could serve as a potential RA diagnostic. VAV1, CD3G, LCK, PTPN6, ITGB2, CXCL13, CD4, and IL7R are found to be previously unclassified, potential biomarkers.

## Introduction

Rheumatoid Arthritis is a clinically heterogeneous and complex autoimmune disease that affects approximately 1.3 million Americans and 20 million people worldwide^14^. The disease manifests between the age of 20 and 40 years, being twice more common in women than men. According to the World Health Organization (WHO), within ten years of the onset of RA, at least 50% of patients in developed countries have to discontinue a full-time job due to the disability that ensues with RA^8^. RA increases inflammation throughout the body, leading to the risk of heart disease and attack on the pericardium. It can also lead to obesity as the joint pain results in decreased exercise among patients^49^. RA is characterized by the release of inflammatory chemicals by the immune system that attacks the synovium, which is the tissue lining around a joint that produces a fluid to help the joint move smoothly. The inflamed synovium thickens and causes the joint area to feel painful and tender. Uncontrolled inflammation can cause destruction and wear down the cartilage, leading to joint deformities and painful bone erosion^37^. Early diagnosis of RA is essential to promote effective treatments. However, it is relatively difficult to do so, since RA’s early-stage symptoms are usually subtle and generic, such as fatigue, slight fever, and stiffness^38^.

Established RA displays clinical heterogeneity, reflected by variable diagnosis leading to the unpredictable, rapid progression of structural damage and inconsistent response to therapy^26^. There have been advancements in synthetic and biological therapies that target specific immune-mediated pathways, but a significant number of patients still fail to respond to current medication--with only 20-30% reaching low disease activity status^26^. The mechanistic reason for such failure rates remains unknown. However, the cellular and molecular variation found in the synovial tissue of patients with long-standing RA is likely to play a role in the variable treatment response and heterogeneous clinical evolution. RA is partially due to genetics, therefore it could potentially be diagnosed through genetic screening, but other cases arise spontaneously or due to environmental factors and autoimmunity^24^.

Early detection and personalized approaches to RA treatment are known to improve patients’ treatment outcomes, minimizing or preventing joint destruction^51^. Therefore, diagnostic tests and biomarkers are in demand for RA. There are various likely avenues for biomarker discovery, and researchers are actively exploring various factors like antibodies and serum. While ACR & EULAR criteria are based mostly on blood tests that measure the ESR and levels of CRP, RF, & ACPAs, such serologic parameters do not necessarily reflect biological actions in the target tissue of the patient and are relatively non-specific to RA^4, 17^. Synovial tissue biomarkers are especially attractive as they can provide a confirmed diagnosis of RA.

Currently, there is still a shortage of an accurate synovial tissue-based biomarker panel for RA diagnosis. Besides diagnosis tests, synovial-tissue-based biomarkers could be applied to develop novel RA treatments, developing therapeutics that can halt or reverse RA progression.

Tissue-based molecular biomarkers can be profiled using various methods, such as microarrays and RNA-seq, both of which are used to perform gene expression studies^42^. Microarray data has been previously used to identify biomarkers for other autoimmune and skeletal system disorders, such as osteoarthritis (OA)^23^. Differential gene expression among RA patients, OA patients, and controls provides detailed information about the mechanisms of RA and could offer specific valuable biomarkers for diagnosis. Studies have previously identified RA biomarker genes through RNA-seq and microarray analysis. For example, one research group identified IL7R as a potential RA biomarker using microarray data^53^. Anti-Cyclic Citrullinated Peptide and Rheumatoid Factor have also been established as serum biomarkers^44^.

Although gene expression methods are instrumental, there are often discrepancies between data due to differences in methodologies or samples, meaning that genes identified as significant in one study might be not so in another. To get greater confidence in biomarkers, it is important to identify genes that are significant across multiple studies, which decreases the likelihood of genes being expressed due to chance.

In this study, the microarray data from 3 separate studies were analyzed to deduce biomarkers for RA. The studies gathered data on gene expression based on the synovial tissue of RA and control patients. DEGs of interest were identified across studies, and their interactions were studied further.

## Methods

### GEO Dataset Selection

NCBI has created a data repository called Gene Expression Omnibus (GEO) that contains public gene expression data from various studies^3, 16^. Datasets matching “synovial,” “Rheumatoid Arthritis,” “expression profiling by array,” and “*Homo sapiens*” were searched in GEO. The GSE55235, GSE2053, and GSE1919 datasets were selected for this study^47, 48, 50^. Through three studies, a total of 19 RA patients and 19 controls were analyzed. All three studies used microarrays for collecting gene expression data from RNA obtained from synovial tissue or synovial membrane was profiled. Microarrays are a chip-based technology that uses probes to capture strands of complementary DNA (cDNA), which is reverse transcribed from the mRNA in the tissue samples, to measure the level of gene expression^20^.

### GEO2R Differential Expression Analysis

GEO provides the GEO2R tool which was used to analyze the three datasets^3, 16^. GEO2R processes gene expression data to identify differentially expressed genes (DEGs) between user-defined groups, in this case, RA and control. For each dataset, thousands of genes were found by GEO2R as statistically significant. T-tests were then used to determine p-values, and GEO2R calculated the fold change for each gene. Fold change is the ratio of the average expression of a gene in one group divided by the average expression in a different group, in this case, comparing RA expression values vs. control values. Some genes are overexpressed in RA while others are underexpressed in RA to controls, hence fold change can be greater than or less than one.

The normal distribution of gene expression values using box-plotting was verified using GEO2R and no outliers were present. Since the gene expression values were all relatively similar, normalization was not required.

Google sheets were used to process the DEGs obtained from GEO2R. DEGs with p-values greater than 0.05 were removed, and the remaining DEGs were then sorted into the two categories of overexpression and underexpression, as some genes were less expressed in RA than the control, while others are more highly expressed in RA. Only genes with a fold change greater than 1.25 were kept.

### Classification of Common Genes

A Venn Diagram for the classification of genes in each of the three studies was generated using the limma package in R studio^39^. Genes found in only one of the three studies were removed from the list. Genes found in only two of the three studies were saved separately from the genes found in all three studies.

### STRING and Cytoscape Network Analysis

The STRING database was used to analyze the two final lists of DEGs, including genes found in all three datasets and the genes found in two out of three datasets. STRING was used to create a network of the protein-protein interactions, and calculate statistically significant Gene Ontology (GO) processes and pathways^43^. A tab-separated value (TSV) file containing the text version of the gene graph was inputted into Cytoscape, a software that can perform more robust analyses and functions on networks^40^. Cytoscpae’s Network Analyzer tool calculated gene network metrics like clustering coefficient and node degree, which is the number of genes connected to a particular gene. When comparing the DEGs found in the list with at least two of the three datasets, hub genes were genes with a node degree of ≥ 8. When comparing the DEGs found in the list with all three datasets, hub genes with a node degree of ≥ 3.

### Gene Ontology Analysis

The biological pathways, processes, and genes linked to them are stored in a database called Gene Ontology (GO) ^2, 19^. The built-in PANTHER tool in GO accepts a gene list and identifies relevant biological mechanisms^29^. Enriched biological processes and molecular functions were isolated from the two DEG lists that were submitted to PANTHER.

### Kegg Analysis

The hub genes identified were inputted into the Kyoto Encyclopedia of Genes and Genomes (KEGG) database to analyze the high-level function of the biological systems that the inputted genes are involved in^22^.

### Reactome Analysis

The genes found in all three datasets were inputted into Reactome to analyze the cellular pathways involved. Reactome is a browser-based database storing information on molecular details of various biological pathways^21^.

## Results

GSE55235, GSE2053, and GSE1919 were analyzed in GEO2R, which generated a list of DEGs. The lists were downloaded, and gene expression value distribution was confirmed to be normal with no outliers based on box-plots of the sample expression distribution.

**Figure 1,2,3:**
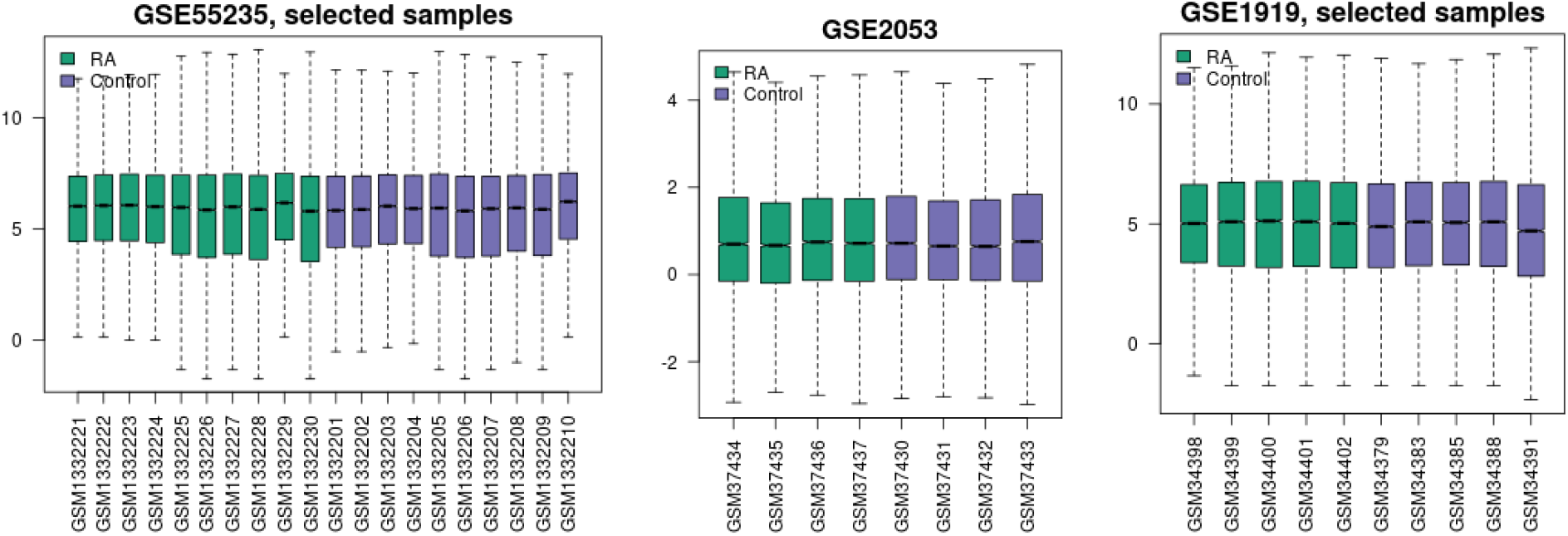
Box plot for gene expression value distribution for GSE55235, GSE2053, and GSE1919. There are no units for the y-axis. The box plots represent the gene expression values for patient samples.

Cut-off criteria for genes were p-value < 0.05. In GSE55235, 5754 genes passed the criteria, in GSE2053 960, and in GSE1919 2220. There was substantial overlap between studies. 64 genes were found in all three datasets, while 1264 genes were found in two out of three datasets.

**Figure 4:**
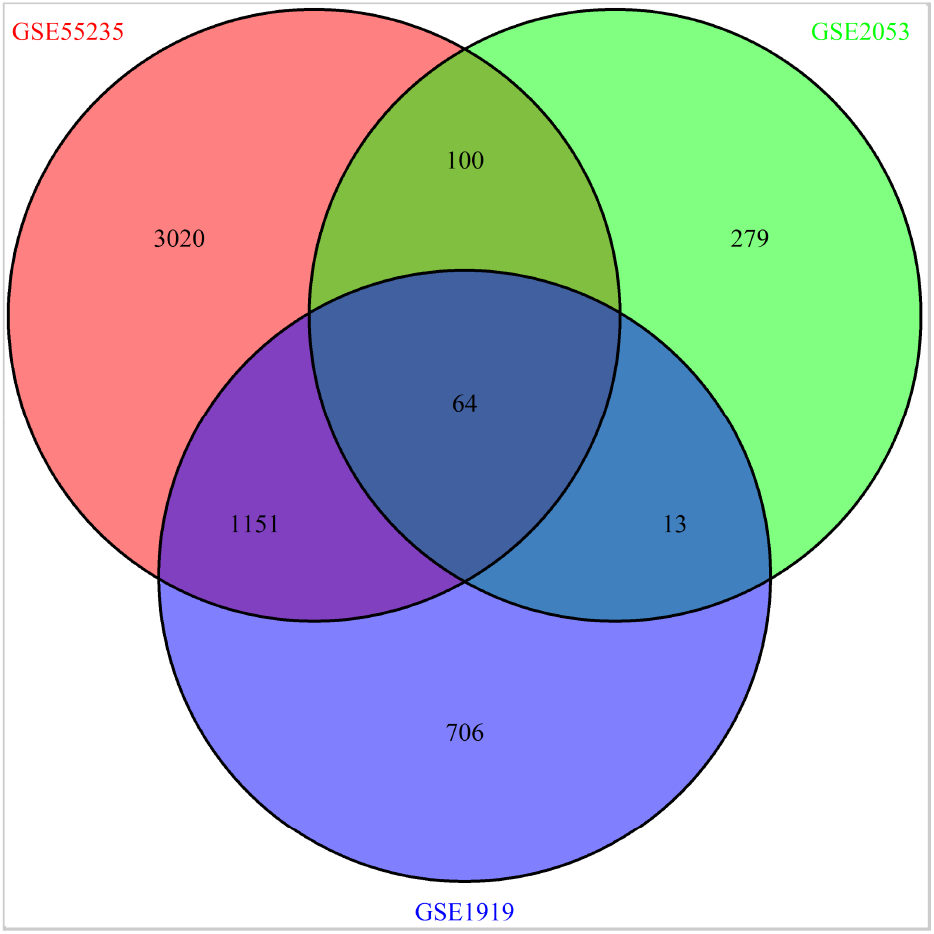
The Venn diagram shows the number of genes that are shared between GSE55235, GSE2053, and GSE1919. 64 genes were differentially expressed in all three sets.

First, the genes that were common in two out of the three microarray dataset were analyzed. Out of the 1264 shared genes in all three datasets, further cut-off criteria were applied: fold change >= 1.25^11^. 320 genes passed all cut-off criteria and were submitted to the STRING tool^43^. 292 out of 320 genes were successfully mapped, as shown in the network below. Solitary genes with no interactions are hidden. The minimum confidence score for protein interactions was set to 0.900 (p-value = 0.009). There were 559 connections between proteins instead of 234 expected interactions, indicating that the network has significantly more interactions than prognosticated of a random sample.

**Figure 5:**
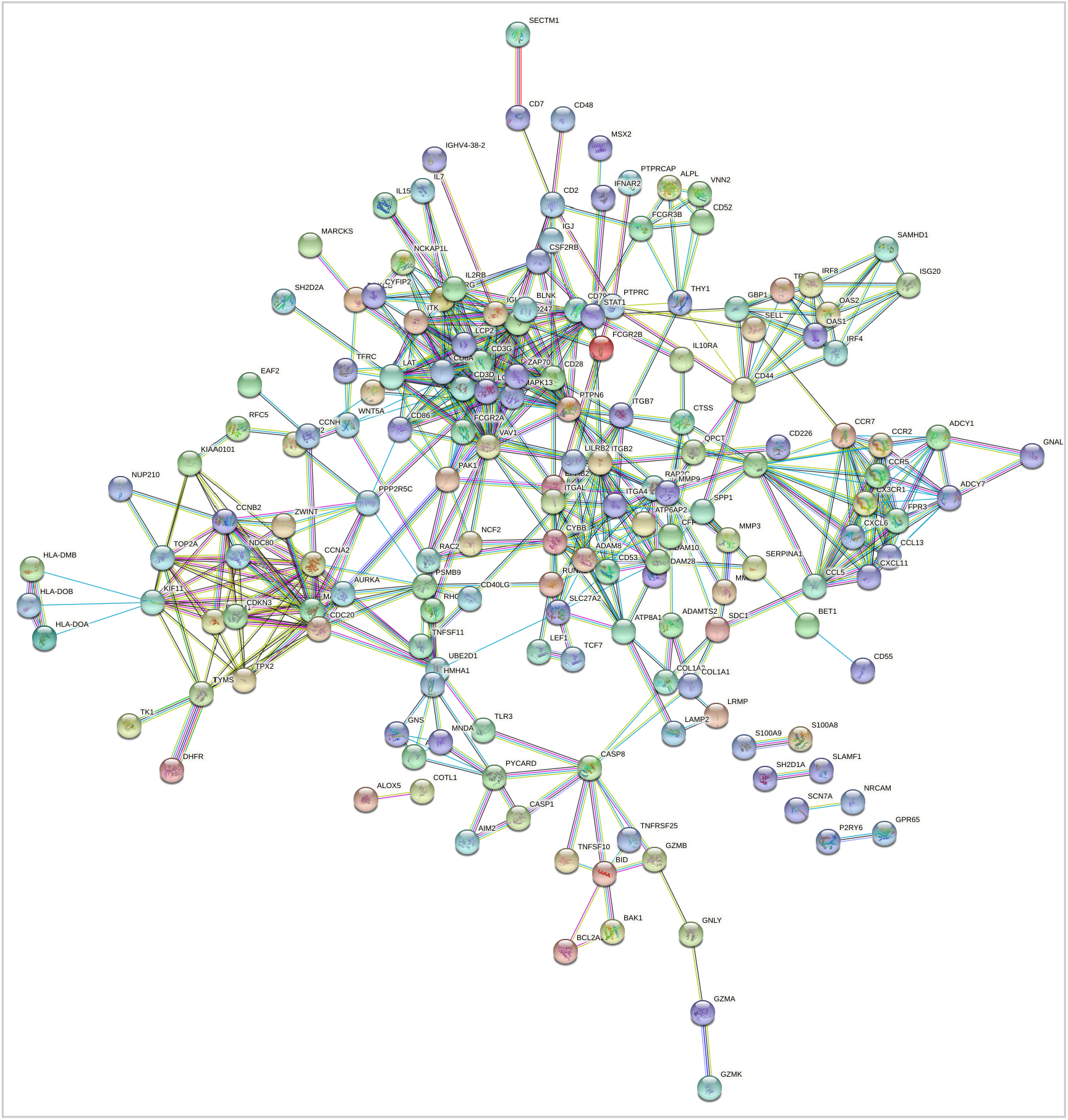
STRING network of protein interactions between 292 DEGs.

The colors of the nodes are arbitrary, and the colors of the edges connecting the nodes display how the interaction was determined. STRING identifies several biological processes and interactions as significant among the inputted genes.

Next, Cytoscape software was used to analyze network characteristics, particularly node degree, to find hub genes.

**Table 1:** Cytoscape generated clustering coefficients and node degrees for the genes in the network. The clustering coefficient measures how clustered nodes are around a single node, specifically the hub genes. A clustering coefficient of 1 represents the maximum density of a node cluster while 0 is the least dense.

| shared name | name | ClusteringCoi | Degree |
| --- | --- | --- | --- |
| VAV1 | VAV1 | 0.310769... | 26 |
| CD3G | CD3G | 0.419913... | 22 |
| LCK | LCK | 0.402597... | 22 |
| PTPN6 | PTPN6 | 0.336842... | 20 |
| ITGB2 | ITGB2 | 0.252631... | 20 |
| CD3D | CD3D | 0.551470... | 17 |
| CDC20 | CDC20 | 0.533333... | 16 |
| CXCL1 | CXCL1 | 0.541666... | 16 |
| CD28 | CD28 | 0.416666... | 16 |
| CD247 | CD247 | 0.625 | 16 |
| CCNA2 | CCNA2 | 0.533333... | 15 |
| CYBB | CYBB | 0.461538... | 14 |
| PAK1 | PAK1 | 0.307692... | 14 |
| CCNB2 | CCNB2 | 0.560439... | 14 |
| KIF11 | KIF11 | 0.527472... | 14 |
| LCP2 | LCP2 | 0.681318... | 14 |
| AURKA | AURKA | 0.560439... | 14 |
| IGLL5 | IGLL5 | 0.340659... | 14 |
| MAD2L1 | MAD2L1 | 0.582417... | 14 |
| ZAP70 | ZAP70 | 0.807692... | 13 |
| IL2RG | IL2RG | 0.423076... | 13 |
| ITGAL | ITGAL | 0.397435... | 13 |
| IL2RB | IL2RB | 0.423076... | 13 |
| CD8A | CD8A | 0.756410... | 13 |

Genes with a node degree of at least 13 are presented, with VAV1, CD3G, LCK, PTPN6, ITGB2, and CD3D with the highest degree.

STRING found 26 gene products that interacted with VAV1. VAV1 codes for an intracellular signal transduction protein involved in T cell proliferation, development, and signal transduction. It has been suggested that VAV1 is involved with rheumatoid arthritis as well as osteogenesis, lymphopoiesis, cardiovascular homeostasis, various cancers, autoimmune diseases, and ML^31^.

The PANTHER tool was then used to analyze the gene lists. PANTHER was set to Annotation Data Set: GO biological process complete, Test Type: Fisher’s Exact, and Correction: Calculate False Discovery Rate (FDR).

PANTHER identified several statistically significant biological processes^45^. Notably, the “negative regulation of antigen processing and presentation of peptide antigen via MHC class II” was enriched by 67.52 times with p-value = 1.25E-03 and an FDR of 3.21E-02. The enrichment value is a number that is the genes in the inputted list that belong to a specific biological process divided by the expected number of genes that would belong to a specific biological process if taken from a random sample of genes. A higher enrichment means more genes in an inputted list are involved in a particular biological process. “Positive regulation of hematopoietic stem cell migration”, “interleukin-10 mediated signaling pathway”, “neutrophil aggregation”, “DN2 thymocyte differentiation”, “cell-matrix adhesion involved in amoeboid cell migration”, and “positive regulation of type III interferon production” were also enriched 67.52 times.

**Table 2:**
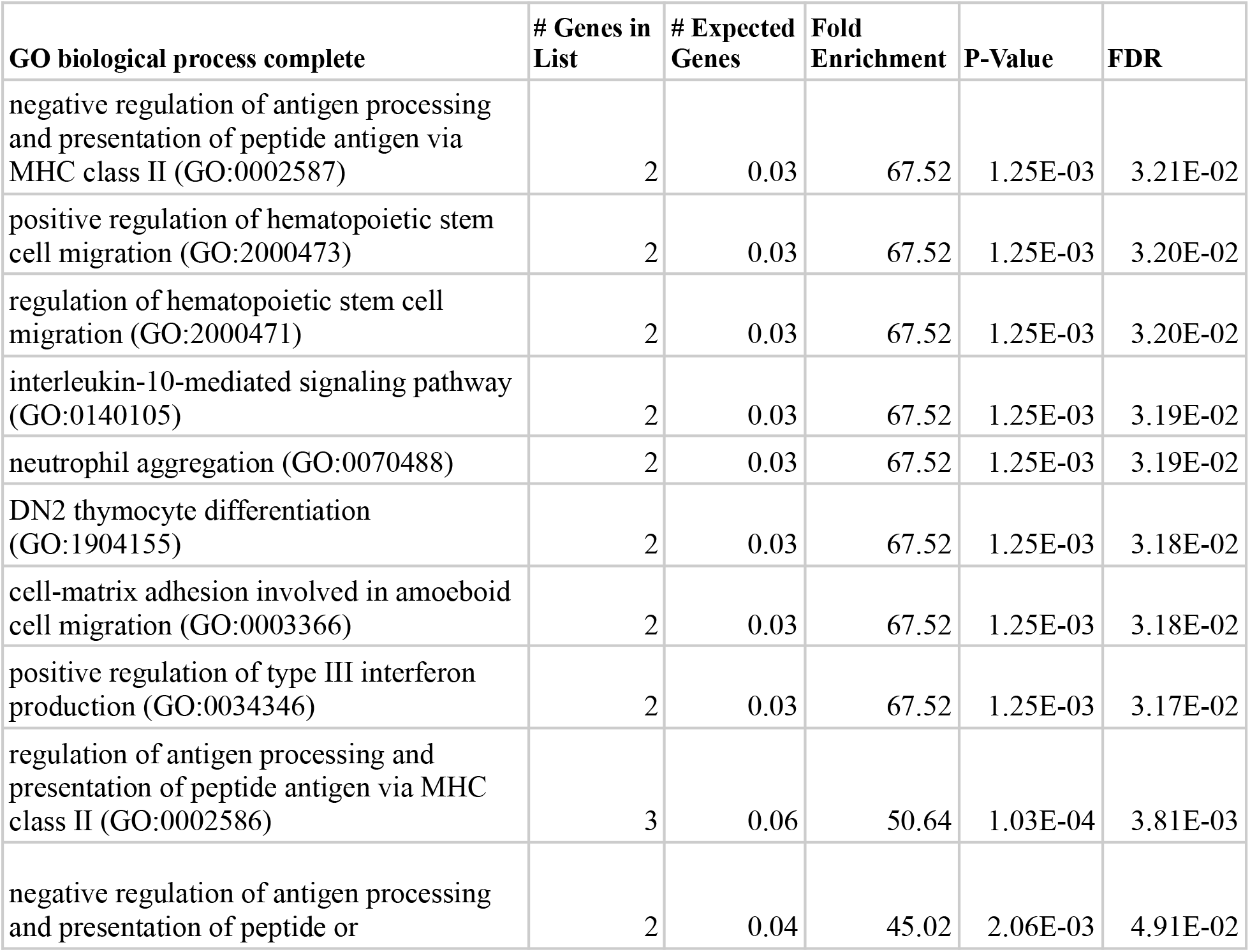
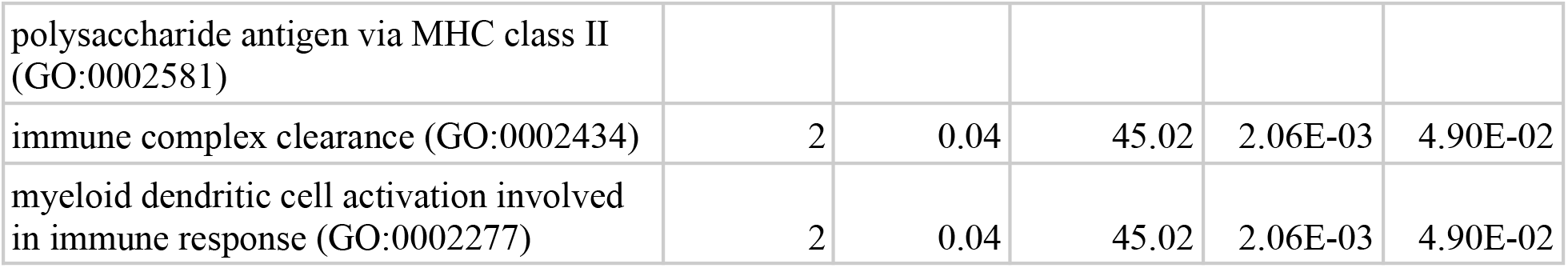
panther outputs a table showing biological processes, ranked by fold enrichment, that are overrepresented or under-represented among the list of genes. GO accession terms are provided for each process.

Many GO molecular functions were also altered. The deoxycytidine deaminase activity function was enriched by 25.32 times with a p-value of 4.66E-04 and an FDR of 4.73E-02. Receptor activity and protein binding appear to be altered in RA.

**Table 3:** panther outputs a table showing the various molecular functions that are over or under-represented among the list of genes. GO accession terms are provided for each function.

| GO molecular function complete | # Genes in List | # Expected Genes | Fold Enrichment | P-Value | FDR |
| --- | --- | --- | --- | --- | --- |
| deoxycytidine deaminase activity (GO:0047844) | 3 | 0.12 | 25.32 | 4.66E-04 | 4.73E-02 |
| transmembrane receptor protein tyrosine phosphatase activity (GO:0005001) | 5 | 0.25 | 19.86 | 1.38E-05 | 2.53E-03 |
| transmembrane receptor protein phosphatase activity (GO:0019198) | 5 | 0.25 | 19.86 | 1.38E-05 | 2.43E-03 |
| C-C chemokine receptor activity (GO:0016493) | 6 | 0.34 | 17.62 | 3.29E-06 | 7.86E-04 |
| C-C chemokine binding (GO:0019957) | 6 | 0.36 | 16.88 | 4.06E-06 | 8.83E-04 |
| MHC class II protein complex binding (GO:0023026) | 4 | 0.24 | 16.88 | 1.79E-04 | 2.04E-02 |
| MHC protein complex binding (GO:0023023) | 6 | 0.37 | 16.21 | 4.98E-06 | 1.03E-03 |
| G protein-coupled chemoattractant receptor activity (GO:0001637) | 6 | 0.39 | 15.58 | 6.05E-06 | 1.21E-03 |
| chemokine receptor activity (GO:0004950) | 6 | 0.39 | 15.58 | 6.05E-06 | 1.16E-03 |
| chemokine binding (GO:0019956) | 7 | 0.49 | 14.32 | 1.62E-06 | 4.31E-04 |
| immunoglobulin binding (GO:0019865) | 5 | 0.36 | 14.07 | 5.71E-05 | 7.57E-03 |

The genes that were only present in all three sets were also analyzed.

Out of the 64 shared genes in all three datasets, further cut-off criteria were applied for stringency: fold change >= 1.25^1^. 24 genes passed all cut-off criteria and were submitted to the STRING tool. 17 out of 24 genes were successfully mapped, and the following graph was created. Solitary genes are hidden to improve clarity. The minimum confidence score for whether a protein interaction existed was set to 0.400 (p-value = 0.004). There were 16 connections between proteins instead of an expected 3, indicating that the network has significantly more interactions than predicted of a random sample.

**Figure 6:**
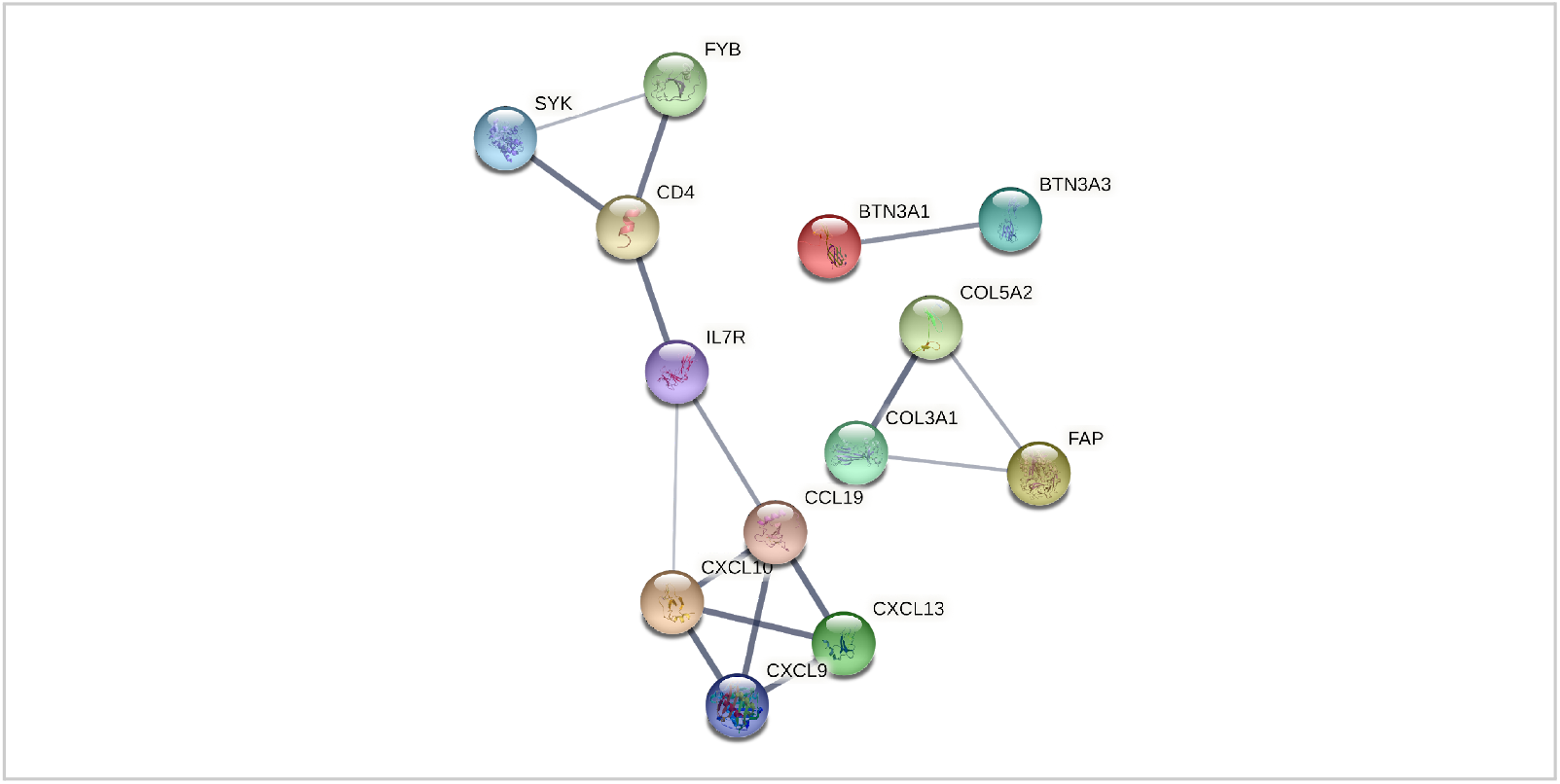
STRING network of protein interactions between 17 DEGs. IL7R, a purple node, is shown to have many interactions with other proteins.

Cytoscape software was then used to analyze the node degree network characteristic to identify hub genes.

**Table 4:** Cytoscape generated clustering coefficients and node degrees for the genes in the network. The genes with a node degree of at least 3 are presented: CXCL13, CXCL9, CXCL10, CCL19. CD4, and IL7R.

| shared name | name | AverageShor | ClusteringCo | ClosenessCei |
| --- | --- | --- | --- | --- |
| CXCL13 | CXCL13 | 2.285714... | 1.0 | 0.4375 |
| CXCL9 | CXCL9 | 2.285714... | 1.0 | 0.4375 |
| CXCL10 | CXCL10 | 1.714285... | 0.666666... | 0.583333... |
| CCL19 | CCL19 | 1.714285... | 0.666666... | 0.583333... |
| CD4 | CD4 | 1.857142... | 0.333333... | 0.538461... |
| IL7R | IL7R | 1.571428... | 0.333333... | 0.636363... |

STRING found 4 gene products to interact with CXCL10. CXCL10 codes for a chemokine and ligand for the receptor CXCR3. Binding of the CXCL10 protein to CXCR3 results in the stimulation of monocytes, natural killer migration, & T-cell migration^7^. This gene has been known to be involved in cytokine storm inflammatory response. CXCL10 was upregulated in all three microarray datasets.

The PANTHER tool was then used to analyze the gene lists. The previously mentioned settings were utilized.

PANTHER identified a number of statistically significant biological processes. The process “regulation of T cell chemotaxis” was enriched >100 times, with p-value 1.36E-04 and FDR of 1.87E-02. “Regulation of myoblast fusion”, “positive regulation of cell adhesion mediated by integrin”, “regulation of lymphocyte chemotaxis”, “regulation of syncytium formation by plasma membrane fusion”, and “positive regulation of cytokine secretion” were also enriched.

**Table 5:** panther outputs a table showing the various biological processes that are over or under-represented among the list of genes.

| GO biological process complete | Genes in List | Expected Genes | Fold Enrichment | P-Value | FDR |
| --- | --- | --- | --- | --- | --- |
| regulation of T cell chemotaxis (GO:0010819) | 2 | 0.02 | > 100 | 1.36E-04 | 1.87E-02 |
| regulation of myoblast fusion (GO:1901739) | 2 | 0.02 | > 100 | 1.86E-04 | 2.38E-02 |
| positive regulation of cell adhesion mediated by integrin (GO:0033630) | 2 | 0.02 | 98.07 | 2.23E-04 | 2.73E-02 |
| regulation of lymphocyte chemotaxis (GO:1901623) | 2 | 0.02 | 82.38 | 3.09E-04 | 3.48E-02 |
| regulation of syncytium formation by plasma membrane fusion (GO:0060142) | 2 | 0.03 | 76.28 | 3.57E-04 | 3.86E-02 |
| positive regulation of cytokine secretion (GO:0050715) | 2 | 0.03 | 73.55 | 3.83E-04 | 4.05E-02 |
| response to vitamin D (GO:0033280) | 2 | 0.03 | 71.02 | 4.09E-04 | 4.27E-02 |
| positive regulation of T cell migration (GO:2000406) | 2 | 0.03 | 68.65 | 4.36E-04 | 4.49E-02 |
| positive regulation of calcium-mediated signaling (GO:0050850) | 2 | 0.03 | 64.36 | 4.92E-04 | 4.79E-02 |
| neutrophil chemotaxis (GO:0030593) | 5 | 0.08 | 64.36 | 1.56E-08 | 4.96E-05 |
| lymphocyte chemotaxis (GO:0048247) | 3 | 0.05 | 63.05 | 1.67E-05 | 3.91E-03 |

Many GO molecular functions were also altered. CCR10, CXCR3, and CXCR chemokine receptor binding functions were enriched by >100 times, which is quite substantial. Receptor binding functions appear to be commonly altered in RA as well.

**Table 6:** panther outputs a table showing the various molecular functions that are over or under-represented among the list of genes.

| GO molecular function complete | Genes in List | Expected Genes | Fold Enrichment | P-Value | FDR |
| --- | --- | --- | --- | --- | --- |
| CCR10 chemokine receptor binding (GO:0031735) | 2 | 0 | > 100 | 8.93E-06 | 6.09E-03 |
| CXCR3 chemokine receptor binding (GO:0048248) | 3 | 0 | > 100 | 4.36E-08 | 1.04E-04 |
| CXCR chemokine receptor binding (GO:0045236) | 3 | 0.02 | > 100 | 1.03E-06 | 9.81E-04 |
| chemokine activity (GO:0008009) | 4 | 0.05 | 85.81 | 1.69E-07 | 2.70E-04 |
| chemokine receptor binding (GO:0042379) | 4 | 0.07 | 60.57 | 6.35E-07 | 7.59E-04 |
| cytokine receptor binding (GO:0005126) | 5 | 0.27 | 18.86 | 5.65E-06 | 4.50E-03 |
| cytokine activity (GO:0005125) | 4 | 0.23 | 17.68 | 7.14E-05 | 4.26E-02 |
| signaling receptor binding (GO:0005102) | 12 | 1.65 | 7.27 | 6.95E-09 | 3.32E-05 |

These genes are then inputted into Reactome to analyze the pathways that were involved^21^. Pathways in the immune system, related to signal transduction, transport of small molecules, gene expression, disease, cell cycle, extracellular matrix organization, and programmed cell death were shown to be involved with the inputted list of genes.

**Figure 7:**
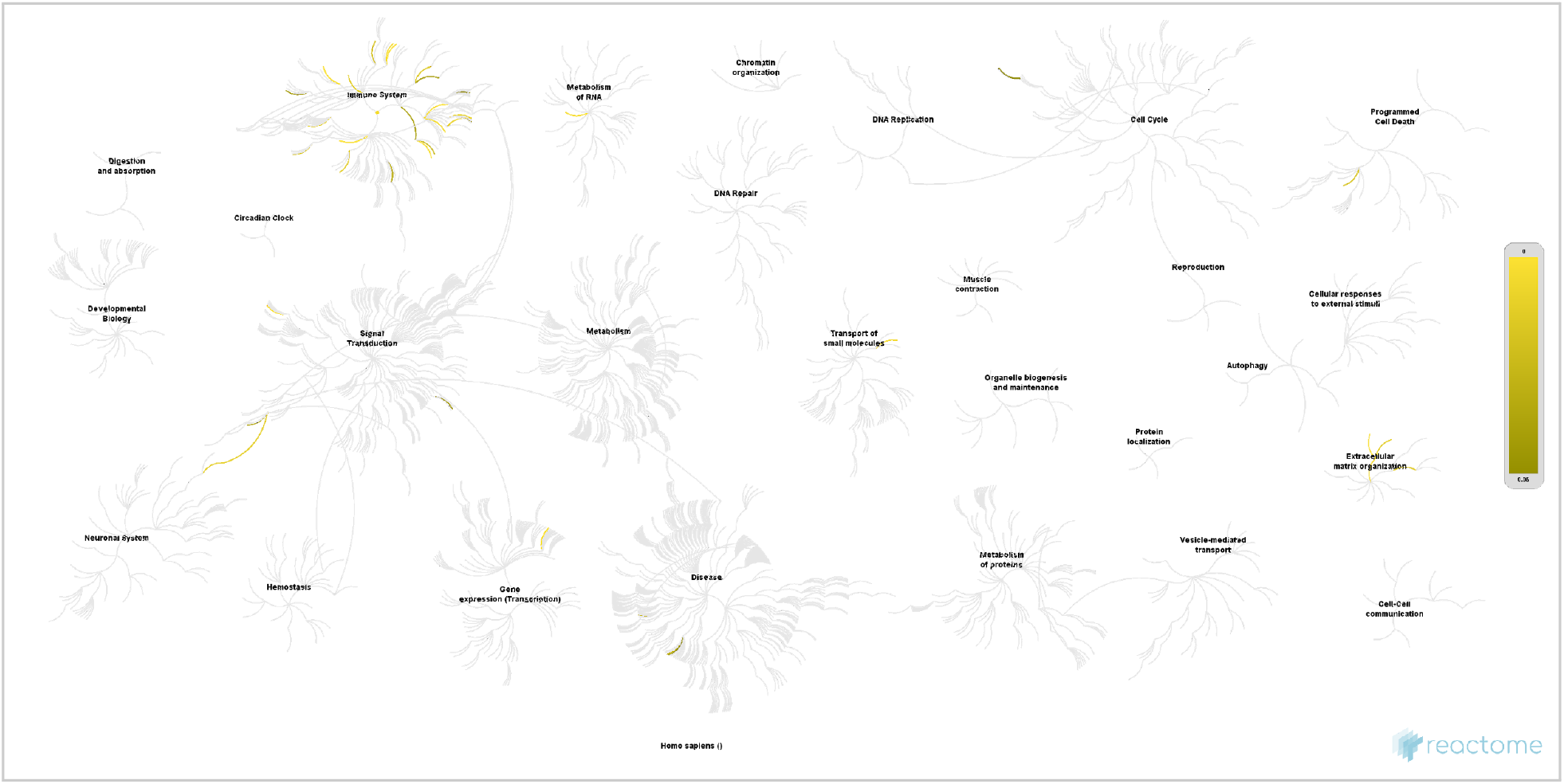
Reactome outputs a map of pathways. Gray pathways represent inputted genes that are not involved in the respective pathways, while yellow-colored pathways are those involved with the inputted genes.

## Discussion

Rheumatoid arthritis is a common autoimmune disease that is often difficult to diagnose at early stages due to the lack of specific symptoms. A synovial tissue-based gene biomarker approach may be beneficial in diagnosing patients earlier to receive more effective treatment.

Analysis of the 3 microarray datasets on RA gene expression yielded 344 DEGs found across studies. These genes pass criteria for p-value and fold change and are attractive for further research as they are both biologically relevant and reproducible. While 320 genes were present in two out of three studies, only 24 were found in all datasets, demonstrating how variable gene expression studies can be and underscoring the need for this study.

VAV1, CD3G, LCK, PTPN6, and ITGB2 had at least 13 interactions with other genes, indicating that they play an essential role in RA. VAV1, LCK, CD3G, and CD3D were located in a cluster along with other genes, such as LAT, CFIP2, CD86, and ZAP70. CXCL13, CXCL9, CXCL10, CCL19, CD4, and IL7R were found in all three datasets and had at least three interactions with other genes, indicating that they may also play an important role in RA. CXCL13, CXCL9, CXCL10, and CCL19 were located in a cluster.

VAV1 was shared across two studies and was a hub gene with degree 26. There is a suggested association between the VAV1 gene rs2617822 polymorphism and RA, yet this research was only preliminary [31]. This study confirms the need for an investigation into the role of VAV1, and other VAV family proteins, in RA pathogenesis. The aforementioned genes that VAV1 was found clustered with, namely LCK, CD3G, CD3D, LAT, CFIP2, CD86, and ZAP70, indicate a potential signal transduction pathway that is useful in RA pathogenesis.

CD3G is a hub gene with degree 22. It is known to play an essential role in the regulation ofT-cell receptor (TCR) expression at the cell surface and is also involved in signal transduction in T-cell activation^12^. Currently, there is interest in elucidating the pathways that drive the production of cytokines and other inflammatory mediators in RA. However, the current kinases that are being researched are ubiquitously expressed, which can lead to side effects^15^. There is a need for more specific targets that can lead to the complete resolution of chronic inflammation. CD3G is not ubiquitously expressed, meaning it can serve as a specific target. Further elucidation on this could lead to the identification of successful therapeutic targets^6^.

**Figure 8:**
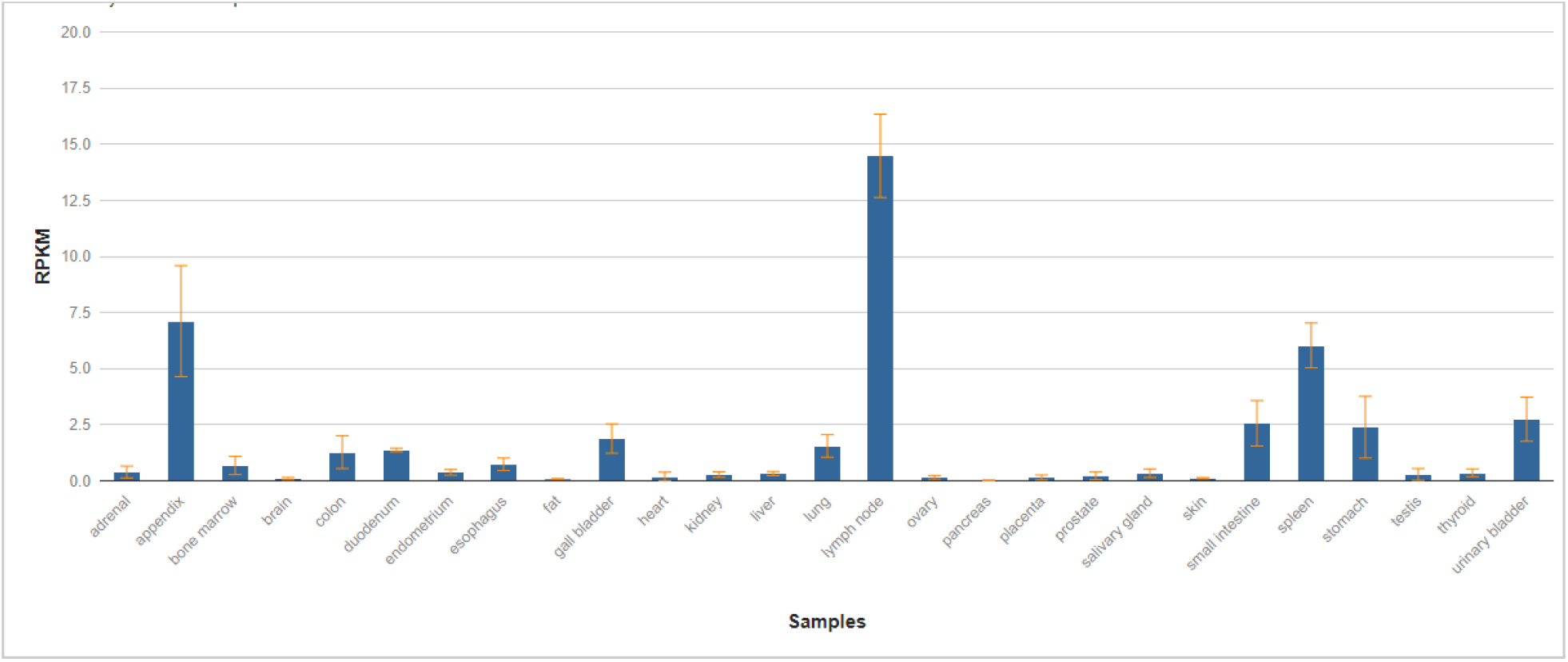
Expression analysis of HPA RNA-seq normal tissues. RNA-sequencing was performed on tissue samples from 95 human individuals representing 27 different tissues to determine the tissue-specificity of the CD3G gene. The CD3G gene is not ubiquitously expressed, indicating that it can serve as a specific therapeutic target.

LCK is a gene that is involved in T-cell signaling. T cells are components of the immune cell infiltrate that have been detected in the joints of RA patients^28^. Since T cell activation is central to mounting the immune response, the inhibition of LOCK blocks T cell activation and suppresses the immune response. While there has been an indication that LCK can serve as an ideal target for the development of novel immunosuppressant agents, extensive experimentation has not been conducted.

PTPN6, also known as SHP-1, modulates signaling through tyrosine-phosphorylated cell surface receptors and plays a role in hematopoiesis^46^. There has currently only been one study that investigates the role of SHP-1 in arthritis, and this study has only been conducted using animals overexpressing this phosphatase^27^. The activation of the SHP-1 coil is considered as a potential treatment of RA.

CXCL13, CXCL9, CXCL10, CCL19, CD4, and IL7R were found in all three datasets and had at least three interactions with other genes, indicating that they may also play an important role in RA. CXCL13, CXCL9, CXCL10, and CCL19 were located in a cluster.

CXCL13, which is involved in B-cell and tumor cell responses, has been found to be important for the diagnosis of early RA, having had a superior performance as compared to RF and anti-CCP, which have been the past biomarkers for RA^1^. Therefore, CXCL13 should be considered a potential biomarker of RA disease activity.

IL-7, the gene that codes for a protein that is a hematopoietic growth factor that is secreted in the bone marrow, and IL-7R, coding for a protein found on the surface of cells, have been linked to RA. IL-7 has been found to have a role in RA angiogenesis^34^. IL-7R should be explored further to explore its relation to RA pathogenesis.

CCL19 has also been found to have a role in RA angiogenesis^33^. CXCL10 signaling has been found to be involved in the pathogenesis of RA^25^. CXCL9 has been found to have increased serum levels when evaluated by ELISA^35^. CXCL9 may serve as a potential biomarker for RA These six genes were inputted into KEGG. As shown in the table below, KEGG indicates that 5 out of the 6 genes are involved in the Cytokine-cytokine receptor interaction pathway. This pathway is involved in the Chemokine signaling pathway, which is associated with leukocyte transendothelial migration and cytokine production. Chemokines have been found to be involved in RA pathogenesis, especially in CCL2 expression, which may contribute to chronic inflammation that is associated with RA^52^. Targeting the Chemokine signaling pathway as well as the cytokine-cytokine receptor interaction pathway as potential avenues of therapeutics for RA should be explored.

**Figure 9:**
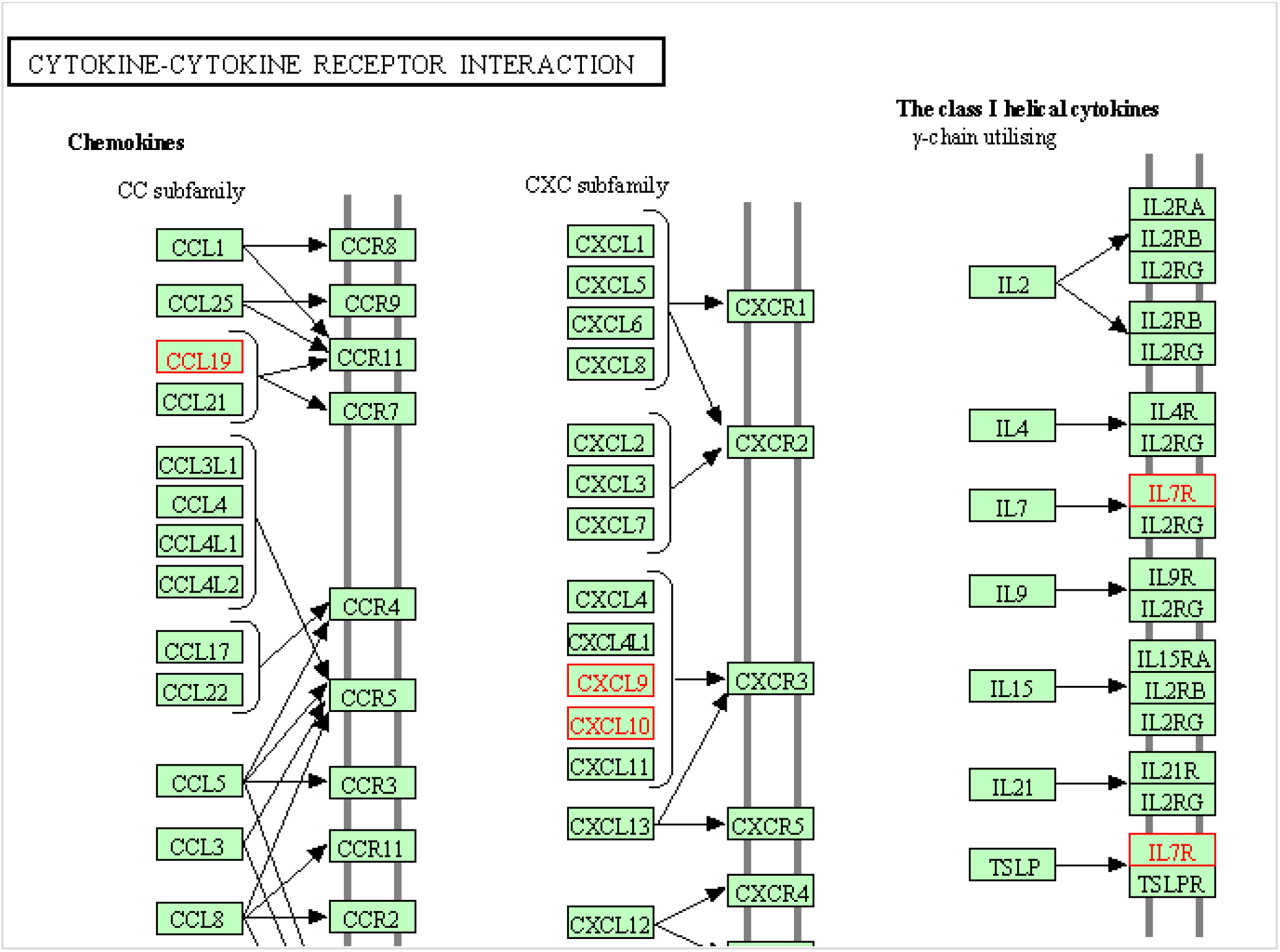
KEGG Mapper Pathway results for CXCL13, CXCL9, CXCL10, CCL19, CD4, and IL7R. 5/6 genes found to be involved in the Cytokine-cytokine receptor interaction pathway - hsa04060. CD4 is not shown in the table as it is in the non-classified section, but it was also found to be interacting in this pathway.

While the hub genes are promising, other genes among the 344 may be of interest as well. “Negative regulation of antigen processing and presentation of peptide antigen via MHC class II” genes are significantly over-represented in the list of DEGs (67.52 times enriched). Peptide bodies have been explored as serum biomarkers^10^. Further research could be done on harnessing the effects of positive stimulation for RA therapeutics.

Genes involved in “positive regulation of hematopoietic stem cell migration” were also overrepresented by 67.52 times. This implies that Hematopoietic stem cells (HSCs), which give rise to other blood cells through Hematopoiesis, migrate to the defined microenvironments in the bone marrow in which they can proliferate and survive^9^. Further research could be done exploring HSCs as an indicator of RA pathogenesis.

“Regulation of T cell chemotaxis” genes were expressed>100 times in the list of DEGs found in all three sets. T cells and other immune cells do migrate to the synovial tissue during RA, which contributes to its pathogenesis^28^. While studies have dashed the promise of targeting individual cytokine receptors of RA, alternative strategies that are aimed at the interactions between the chemokine signaling pathways presented by KEGG and the other involved genes from all three datasets should be considered for RA therapy.

Reactome presented TICAM-1 deficiency in RA. The TICAM 1 gene codes for a protein that mediates protein-protein interactions between toll-like receptors (TLRs) and signal transduction components^13^. While Toll-like receptors have potentially been linked to RA, not much has been found in TICAM-1 deficiency in RA.

There was also an overrepresentation of the Adenylate cyclase inhibitory pathway. In this pathway, Adenylate cyclase is inhibited, the production of cAMP from ATP is inhibited, which ultimately decreases the activity of cAMP-dependent protein kinase^21^. Becker et al. found that the caAMP pathway can serve as a therapeutic target in autoimmune and inflammatory diseases, as cAMP (Cyclic adenosine monophosphate) is a regulator of innate and adaptive immune cell function^36^. There is a plethora of evidence that suggests that the activation of the innate immune system is linked to RA. cAMP as a therapeutic target is a potential avenue that could lead to better therapeutic development for RA.

Defective SFTP2 causing IPF was also overrepresented in Reactome. Mutations in SFTPA2 disrupt protein structure, and the defective protein is retained in the ER membrane which causes idiopathic pulmonary fibrosis^21^. While formerly RA has been linked with pulmonary fibrosis, this study links RA to the development of IPF.

The strengths of this study include the rigor applied to DEG selection. To qualify for the final list, a gene had to have a fold change of at least 1.25, a p-value of < 0.05, and be present in at least 2 datasets. The selective analysis was done on genes found in all 3 datasets. When inputted into STRING, the highest confidence interval of 0.900 was used to analyze the genes that were in at least 2 out of 3 datasets, indicating the statistical significance of the protein-protein interactions that were identified. Additionally, the samples were mixed between males and females, indicating that the significant differences in gene expression and RA pathology between sexes were taken into account in this study. Another strength is the sample size. RA is known for its clinical heterogeneity between patients. By considering 20 RA patients, there is greater assurance of obtaining a comprehensive view of the gene expression of RA than with just a few samples.

The limitations of this study are that no normalization was performed between datasets, though this is common with cross-study gene expression analysis. Additionally, RA patients were not compared with other arthritis controls. Thus, there is a degree of uncertainty around whether the DEGs found are specific to RA or also occur in, say, osteoarthritis. Yet, considering that RA and osteoarthritis differ as A involves the immune system, there can be a differentiation between the gene expression data. The studies that developed the three datasets also contain gene expression data from osteoarthritis, so the next step in the research is to evaluate the biomarkers found in this study and examine how their expression compares in RA vs. other skeletal disorders. Lastly, the results of this study should be confirmed in vitro to certify that the aforementioned pathways and hub genes are statistically significant.

The DEGs identified are useful as diagnostic markers for RA tests and may also be useful targets for novel treatments for RA. The present study elucidates several pathomechanisms, such as the negative regulation of antigen processing and presentation of peptide antigen via MHC class II, the positive regulation of hematopoietic stem cell migration, and the regulation of T cell chemotaxis, that could be avenues for research in alleviating RA symptoms. Currently, not much is known about the involvement of Hematopoietic stem cells in RA development, however, there is growing interest. Mills et al. find that RA can cause hematopoietic stem cell reprogramming^30^. This study’s finding that HSCs are affected through RA pathogenesis provides further evidence for Mills’s findings.

Future research directions include finding genes that can distinguish between RA and other skeletal and immune conditions. Additionally, epigenetic factors such as miRNAs, histone modification, and PTMs and their interactions with genes should be explored. Examining the role of Hematopoiesis and signal transduction pathways in RA may also yield important information.

## Data Availability

Supplementary information is available for this paper. All sources are cited, and the repository of differentially expressed genes is provided. Correspondence and requests for materials should be addressed to.

https://docs.google.com/spreadsheets/d/11yDBMAnymuDmIV6B3Tc2y1ltM_YRYu_QE04G4mHFQHQ/edit#gid=0

## Supplement

Differentially Expressed Genes https://docs.google.com/spreadsheets/d/11yDBMAnymuDmIV6B3Tc2y1ltM_YRYu_QE04G4mHFQHQ/edit?usp=sharing

